# Leveraging Self-Supervised Learning for Non-Invasive Intra-Cardiac Magnetic Resonance Oximetry Assessment

**DOI:** 10.64898/2026.06.29.26356860

**Authors:** Jiayuan Chen, Thai-Hoang Pham, Ping Zhang, Juliet Varghese

**Affiliations:** Department of Computer Science and Engineering, The Ohio State University, Columbus, Ohio, USA; Department of Biomedical Informatics, The Ohio State University, Columbus, Ohio, USA; Department of Radiology, The Ohio State University, Columbus, Ohio, USA

## Abstract

Accurate measurement of intra-cardiac blood oxygen (O2) saturation is essential for cardiovascular assessment, yet current methods require invasive catheterization. T2-based cardiac magnetic resonance imaging (CMRI) enables non-invasive O2 quantification, but deep learning automation is constrained by scarce annotated data. We propose a unified self-supervised learning (SSL) framework integrating cine CMRI and T2 oximetry CMRI to learn generalizable representations without labels. Our approach pre-trains ResNet and vision transformer encoders using contrastive learning and masked image modeling on over 48,000 cardiac images. Pre-trained encoders are fine-tuned for O2 saturation regression with uncertainty quantification to enhance clinical trustworthiness. Our SSL framework significantly outperforms traditional radiomics and supervised baselines, with SimCLR pre-trained ResNet achieving a mean absolute error of 3.70, representing over 15% improvement. These findings demonstrate SSL’s potential to address annotation bottlenecks in non-invasive cardiac diagnostics.

## 1 Introduction

In recent years, deep learning has achieved remarkable progress in medical image analysis, offering expert-level performance across diverse modalities such as chest X-rays [1, 2], fundus photographs [3, 4], and magnetic resonance imaging (MRI) [5, 6]. Among these, cardiac MRI (CMRI) stands out as a non-invasive, high-resolution modality that plays a central role in evaluating cardiac structure, function and myocardial tissue characteristics [7]. Widely used in clinical practice, CMRI enables diagnosis and monitoring of conditions including cardiomyopathies, ischemic heart disease, and myocarditis [8], with deep learning approaches showing particular promise for automating and enhancing these diagnostic tasks [6, 9]. Beyond structural and functional assessment, there is growing interest in using CMRI to estimate physiological biomarkers that traditionally require invasive measurement.

Building on these advances, recent research has focused on predicting intra-cardiac blood oxygen (O2) saturation from CMRI, a physiological biomarker essential for evaluating cardiovascular conditions such as heart failure and congenital heart disease [10, 11]. Conventional assessment requires invasive catheterization, motivating the development of non-invasive alternatives. T2-based MR oximetry exploits the fact that blood T2 relaxation times are sensitive to O2 saturation, as deoxygenated blood becomes increasingly paramagnetic, thereby enabling non-invasive quantification of O2 levels in the cardiac chambers. However, deriving accurate O2 saturation estimates from T2-weighted images requires robust mapping of image features to physiological values, a task well suited to deep learning. Deep learning models, including convolutional neural networks such as ResNet [12] and transformer-based models such as Vision Transformers (ViT)[13], typically initialized with large-scale supervised pretraining (e.g., ImageNet [14]) and fine-tuned on task-specific data [15], provide a powerful framework for learning complex relationships between cardiac MR image patterns and physiological measurements such as O2 saturation. However, training such models remains challenging because supervised learning relies on abundant labeled data. In the case of intra-cardiac O2 saturation, ground-truth labels are inherently scarce, as their acquisition requires invasive catheterization, fundamentally limiting the feasibility of large-scale supervised training.

To address this annotation bottleneck, we turn to self-supervised learning (SSL), which enables models to learn meaningful representations from unlabeled medical images through surrogate tasks such as contrastive learning [16] and masked image modeling [17]. SSL has demonstrated particular advantages in medical imaging by leveraging vast repositories of unlabeled data and showing strong transferability across different modalities, aligning with the foundation model paradigm of learning universal representations that can be adapted to downstream tasks. In this work, we propose a unified SSL framework that integrates cine and T2 oximetry CMR images to learn generalizable image representations. We leverage large-scale open-source cine MRI data, including the Sunnybrook Cardiac Data (SCD) [18] and Automated Cardiac Diagnosis Challenge (ACDC) [9], and small-scale in-house T2 oximetry MR images, constructing over 48,000 2D cardiac images for self-supervised pre-training. The pre-trained encoders are fine-tuned for prediction of blood oxygen saturation in the right ventricle, with uncertainty quantification to enhance clinical confidence in the estimated O2 saturation. Our results demonstrate significant improvements over both conventional feature extraction methods using radiomics and supervised deep learning baselines, highlighting the value of SSL for cardiac physiological measurement tasks.

## 2 Related Work

### 2.1 T2-based MR Oximetry

T2-based MR oximetry has been investigated as a non-invasive surrogate for catheter-based oxygen saturation measurement, exploiting the sensitivity of blood T2 relaxation times to deoxyhemoglobin concentration [19, 20]. Several studies have demonstrated promising agreement with invasive catheter references [11, 21]. However, existing approaches typically rely on model-based parameter fitting with scanner- or protocol-specific calibration curves, often requiring complex post-processing pipelines and specialized expertise, which has limited broader clinical adoption [22]. An alternative non-invasive approach based on quantitative susceptibility mapping (QSM) has also been explored, exploiting the linear dependence of blood magnetic susceptibility on deoxyhemoglobin concentration to estimate differential oxygen saturation between left and right heart chambers [23]. While cardiac QSM has demonstrated feasibility in healthy volunteers and initial patient validation against catheterization, it remains challenged by background field effects from air in the lungs, chemical shift artifacts from epicardial fat, and cardiac and respiratory motion, limiting its robustness for routine clinical use [24]. These collective limitations of both T2-based and susceptibility-based MR oximetry motivate the development of data-driven approaches that can learn the complex mapping from MR image features to physiological oxygen saturation without reliance on simplified physical models.

### 2.2 Deep Learning for Cardiac MRI

Radiomics-based approaches [25] extract hand-crafted intensity and texture features from cardiac MR images but depend heavily on manual feature engineering and may fail to capture subtle physiological patterns associated with blood oxygenation. Deep learning has been widely applied to cardiac MRI for tasks such as segmentation and disease classification [6, 9], yet these efforts have predominantly targeted structural or morphological endpoints. The application of deep learning to physiological parameter estimation, particularly intra-cardiac oxygen saturation, remains limited, largely because supervised training requires labeled datasets that are inherently scarce when ground-truth acquisition demands invasive catheterization.

### 2.3 Self-Supervised Learning in Medical Imaging

SSL has emerged as a powerful paradigm for learning representations from unlabeled data through surrogate tasks such as contrastive learning [26] and masked image modeling [17]. In the medical imaging domain, SSL has shown strong transferability across modalities and improved performance under limited supervision [27, 28, 29]. Within cardiac imaging specifically, SSL offers the potential to leverage large repositories of cine MRI data to learn anatomy- and motion-aware representations that generalize to downstream tasks [30]. Despite these advances, the application of SSL to non-invasive MR oximetry remains largely unexplored, leaving open the question of whether self-supervised pre-training can improve physiological prediction from cardiac MRI under scarce label conditions.

## 3 Method

Our framework consists of two main stages, as shown in Figure 1: (1) self-supervised pre-training using SimCLR and MAE on large-scale cardiac MR cine data to learn generalizable representations, and (2) supervised fine-tuning for prediction of blood O2 saturation with uncertainty quantification.

**Figure 1:**
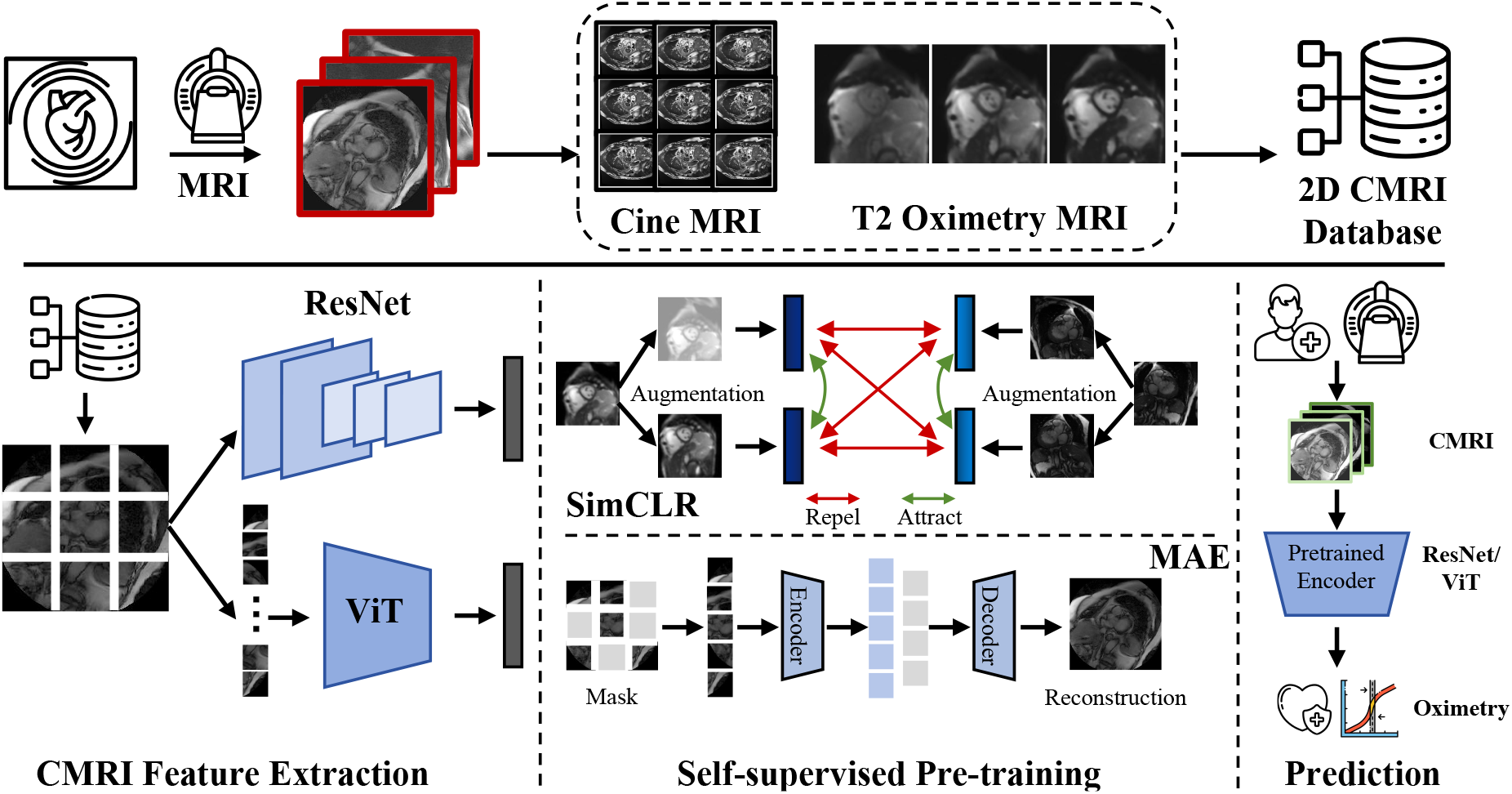
Overview of proposed framework for prediction of intra-cardiac blood oxygen saturation. A 2D CMRI database is collected from cine MRI and T2 sensitive MR oximetry images. Self-supervised pre-training is then performed on commonly used image encoders including ResNet and ViT using SimCLR contrastive learning and masked autoencoder. Subsequently, supervised fine-tuning is conducted on labeled oximetry MR data for prediction of oxygen saturation.

### 3.1 Data Curation

Our dataset is designed to support both self-supervised pre-training on large-scale unlabeled cardiac MRI and supervised fine-tuning for intra-cardiac oxygen saturation estimation. It comprises cardiac MR images acquired in the short-axis plane, including cine MRI and T2-weighted images acquired for MR oximetry. For T2-based oximetry, six T2-prepared images were acquired per patient using a T2-prepared SSFP sequence with varying inter-echo spacings (*τ*_180_ = 0, 5, 10, 15, 20, and 25 ms), providing multi-contrast input with graded sensitivity to blood O2 saturation. The in-house MR oximetry cohort consists of patients who underwent clinically indicated right heart catheterization for hemodynamic evaluation, such as suspected pulmonary hypertension or congenital heart disease assessment. Catheter-derived O2 saturation measurements in the right ventricle serve as ground-truth labels, representing a clinically validated reference obtained as part of routine care rather than an additional research procedure. The cohort was collected under institutional oversight with appropriate approvals/waivers, and all analyses were performed on de-identified data. Cases were excluded if they presented severe motion or susceptibility artifacts, incomplete T2-prepared acquisitions, or absent catheter-based O2 saturation records. To complement the limited labeled oximetry data, we integrate large-scale public cine MRI datasets, including SCD and ACDC, for self-supervised pre-training.

### 3.2 CMRI Feature Extraction

We employ two representative encoder architectures: ResNet-50, a convolutional network with strong local feature extraction, and ViT-Base, a transformer model that captures long-range spatial dependencies. The use of both architectures allows us to compare the effect of different inductive biases on physiological parameter prediction from cardiac MRI. Given a set of *k* T2 sensitive images [*x*_1_, *x*_2_, …, *x*_*k*_] for each patient, we apply the encoder, denoted as Enc, to each image individually to obtain feature embeddings [*h*_1_, *h*_2_, …, *h*_*k*_]. A global representation *h* for patient *i* is then obtained by applying either max pooling or average pooling across the sequence:

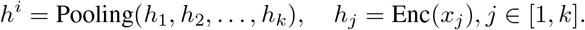

We evaluate both max pooling and average pooling as aggregation strategies. This multi-image aggregation reflects the clinical T2 oximetry protocol, in which multiple images with varying T2 preparation times are acquired per patient; pooling yields a stable patient-level representation that is robust to single-frame artifacts.

### 3.3 Self-supervised Pre-training

To learn generalizable cardiac representations without labeled data, we employ two complementary self-supervised frameworks: SimCLR for contrastive learning and MAE for masked image modeling. SimCLR is architecture-agnostic and is applied to both ResNet-50 and ViT-Base, while MAE is inherently designed for the vision transformer architecture and is therefore applied exclusively to ViT-Base. This setup allows us to compare discriminative and reconstructive pre-training strategies, as well as the effect of architecture choice within the contrastive learning paradigm.

SimCLR uses contrastive learning to maximize similarity between augmented views of the same image while contrasting with negative examples, with the objective:

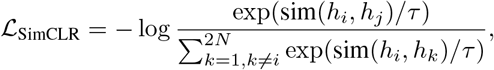

where *h*_*i*_ and *h*_*j*_ are feature representations of augmented views from the same image, *τ* is a temperature parameter, and sim(*·, ·*) denotes cosine similarity.

MAE learns through masked image reconstruction, minimizing the mean squared error between original and reconstructed patches: 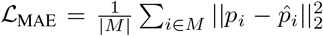, where *M* represents the set of masked patches, *p*_*i*_ is the original patch, and 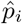 is the reconstructed patch. These approaches enable learning of generalizable cardiac representations without requiring labeled data.

### 3.4 Supervised Fine-tuning with Uncertainty Quantification

Following self-supervised pre-training, the encoder weights are used to initialize the downstream supervised task. The regression head, implemented as a two-layer MLP, is appended to the encoder and outputs both predicted mean *ŷ*_*I*_ and log-variance 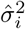, modeling predictions as Gaussian distributions 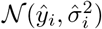. We optimize using Gaussian Negative Log-Likelihood loss:

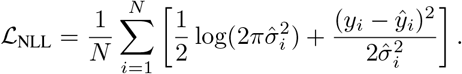

#### Clinical Interpretation

The predicted variance 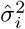 provides a per-patient confidence indicator that can inform clinical decision-making. Cases with elevated uncertainty may reflect challenging imaging conditions, atypical cardiac anatomy, or physiological profiles underrepresented in the training data. In practice, such uncertainty flags could prompt clinicians to seek additional diagnostic confirmation, such as invasive catheterization, rather than relying solely on the model’s point estimate.

#### Classification Task

In addition to continuous regression, we evaluate a binary classification variant that determines whether blood O2 saturation falls within expected normal physiological ranges in the right ventricle (60-80%) [31]. This formulation addresses a clinically actionable screening question: identifying patients with potentially abnormal oxygenation who may require further invasive assessment. The classification head applies a sigmoid activation optimized with binary cross-entropy loss.

## 4 Experiments

In this section, we present comprehensive experiments to evaluate the effectiveness of our self-supervised learning framework for prediction of intra-cardiac blood O2 saturation from T2 prepared MRI data.

### 4.1 Experimental Setup

#### Data

We utilize three CMRI data sources: SCD, ACDC that contain cine MRI images, and our in-house sparse T2-prepared cardiac MR acquired following previously described MR oximetry protocol [11]. The combined dataset includes 48,896 short-axis 2D CMRI images from 252 patients for self-supervised pre-training and supervised fine-tuning. The detailed statistics are shown in Table 1. Public cine datasets used for SSL pretraining were disjoint from the in-house oximetry cohort at the patient level. For clarity, no ground-truth oxygen saturation labels from the in-house cohort were used during the self-supervised pretraining stage. SSL was performed solely using image-level augmentation objectives without access to patient-level physiological measurements. This design ensures that no label information from downstream patients was introduced during representation learning, thereby avoiding data leakage.

**Table 1:** Summary statistics of the three datasets.

| Dataset | SCD | ACDC | SCMRI | All |
| --- | --- | --- | --- | --- |
| # Patient | 45 | 150 | 57 | 252 |
| # 2D Image | 10,178 | 38,346 | 372 | 48,896 |

The labeled oximetry cohort consists of 57 patients with catheter-based oxygen saturation measurements ranging from 49% to 87%. Given the relatively limited cohort size, we analyzed the distribution of oxygen saturation values to characterize potential imbalance across physiological ranges. Figure 2 illustrates the distribution across predefined saturation intervals, highlighting the number of patients within each range.

**Figure 2:**
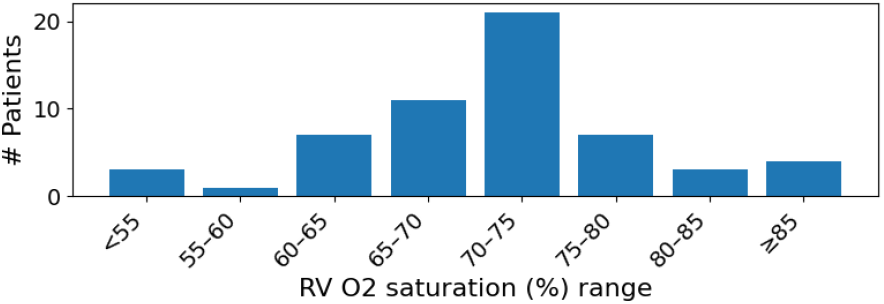
Distribution of catheter-based RV O2 saturation labels.

#### Experimental Setting

Images are resized to 256×256 pixels with random/center cropping to 224×224 for training/evaluation. We evaluate using MAE and RMSE for regression, and Accuracy and F1-Score for binary classification. For the binary classification task (normal range: 60–80%), the cohort included 46 normal cases and 11 abnormal cases; class proportions were preserved across patient-level folds. This class imbalance reflects the distribution of clinically acquired catheter measurements in our cohort.

All experiments were conducted using patient-level splits to prevent information leakage across training and evaluation sets. Given the limited cohort size, we performed five-fold cross-validation at the patient level. In each fold, approximately 80% of patients were used for training and 20% for testing, with no patient overlap across folds. A small validation subset was selected from the training fold for early stopping and hyperparameter tuning. We report the mean and standard deviation across the five folds to provide a robust estimate of model performance.

### 4.2 Implementation Details

We employ ResNet-50 [12] and ViT-Base [13] as backbone architectures. **Pre-training:** ViT uses both MAE (75% masking ratio) and SimCLR, while ResNet uses SimCLR with batch size 512. Pre-training runs for 400 epochs with learning rate 1e-3, cosine scheduling with warmup. Data augmentation includes random flipping, Gaussian blur, and cropping. **Fine-tuning:** Pre-trained encoders are fine-tuned on labeled data using a linear regression head outputting mean and log-variance for uncertainty quantification. We apply 0.1 dropout and train for 100 epochs with early stopping. Experiments use NVIDIA A100 GPUs with PyTorch 2.1. For the radiomics baseline method (Radiomics), we extract hand-crafted radiomics features using PyRadiomics [25] from whole heart regions, including first-order statistics, shape descriptors, and texture features (GLCM, GLRLM, GLSZM). These features are then fed into a linear regressor trained on the same labeled data splits for comparison.

### 4.3 Main Results

The experimental results are presented in Table 2, where Radiomics represents a traditional machine learning method and is used as a baseline to evaluate performance of the proposed models. ResNet and ViT baselines are models fine-tuned from ImageNet pre-trained weights. SimCLR-ResNet represents ResNet pre-trained with SimCLR contrastive learning on our curated cardiac MRI dataset. MAE-ViT and SimCLR-ViT represent ViT models pre-trained using Masked Autoencoder and SimCLR frameworks respectively on the same dataset.

**Table 2:** Comparison of supervised baselines (ResNet and ViT) and self-supervised pre-training methods (SimCLR-ResNet, SimCLR-ViT and MAE-ViT) for prediction of intra-cardiac blood oxygen saturation. Results show mean and standard deviation across five random splits. Best results are bold.

| Task | Metric | Radiomics | ResNet | ViT | SimCLR-ResNet | SimCLR-ViT | MAE-ViT |
| --- | --- | --- | --- | --- | --- | --- | --- |
| Regression | RMSE ↓ | $8.50 \pm 0.15$ | $6.23 \pm 0.32$ | $7.85 \pm 0.09$ | **$5.52 \pm 0.24$** | $5.63 \pm 0.05$ | $5.60 \pm 0.14$ |
| | MAE ↓ | $5.20 \pm 0.08$ | $4.31 \pm 0.02$ | $6.18 \pm 0.26$ | **$3.70 \pm 0.06$** | $3.79 \pm 0.29$ | $3.79 \pm 0.09$ |
| Classification | Acc. ↑ | — | $0.68 \pm 0.06$ | $0.68 \pm 0.06$ | $0.75 \pm 0.05$ | $0.77 \pm 0.06$ | **$0.78 \pm 0.02$** |
| | F1 ↑ | — | $0.81 \pm 0.04$ | $0.81 \pm 0.04$ | $0.85 \pm 0.03$ | $0.87 \pm 0.04$ | **$0.87 \pm 0.01$** |

For the regression task, SimCLR-ResNet achieves the best performance with RMSE of 5.52 and MAE of 3.70, representing substantial improvements of more than 10% respectively compared to the supervised ResNet baseline. MAE-ViT demonstrates the strongest performance among Vision Transformer variants, with RMSE of 5.60 and MAE of 3.79. Notably, the MAE of 3.70 achieved by SimCLR-ResNet approaches the reported measurement variability of invasive catheterization itself (2–5%) [11], suggesting that the model’s prediction error is approaching the noise floor of the reference standard. For the classification task, MAE-ViT attains the highest accuracy of 0.78 and F1-score of 0.87. To further evaluate performance under class imbalance, we report the precision–recall curve of the best-performing MAE-ViT model (Figure 3), achieving an average precision (AP) of 0.80. The curve remains substantially above the positive-class baseline, indicating strong discriminative ability for detecting abnormal oxygen saturation cases. All self-supervised pre-training methods consistently outperformed their corresponding supervised baselines across both tasks. The improvements are particularly pronounced when compared against the supervised ViT baseline, where all SSL variants show substantial gains on both regression and classification tasks. These results demonstrate the effectiveness of leveraging large-scale unlabeled cardiac MR data through self-supervised learning, particularly in scenarios where labeled data for specialized physiological measurements is limited.

**Figure 3:**
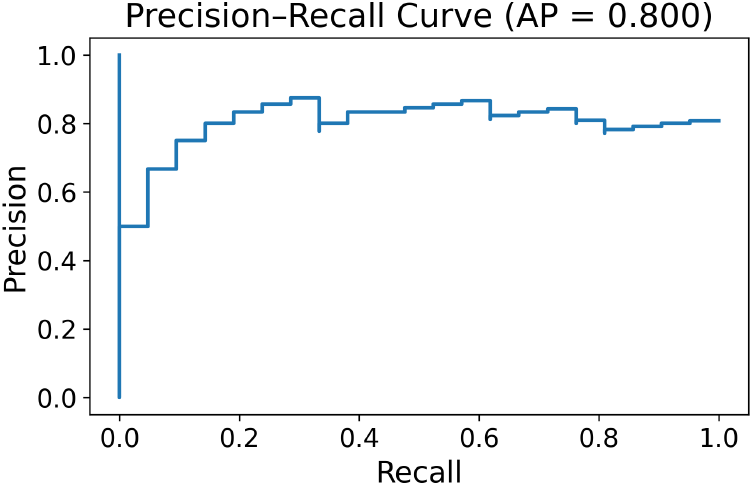
Precision–Recall curve of MAE-ViT.

### 4.4 Model Analysis

#### 4.4.1 Masked Autoencoder Reconstruction Analysis

To qualitatively examine the representations learned during self-supervised pretraining, we visualize MAE reconstructions (Figure 4). The reconstructed images preserve key cardiac anatomical structures, including ventricular geometry and myocardial boundaries, despite substantial masking. Notably, the blood pool region remains well-defined, suggesting that the encoder captures spatial patterns relevant to ventricular cavity characterization rather than relying on superficial texture cues.

**Figure 4:**
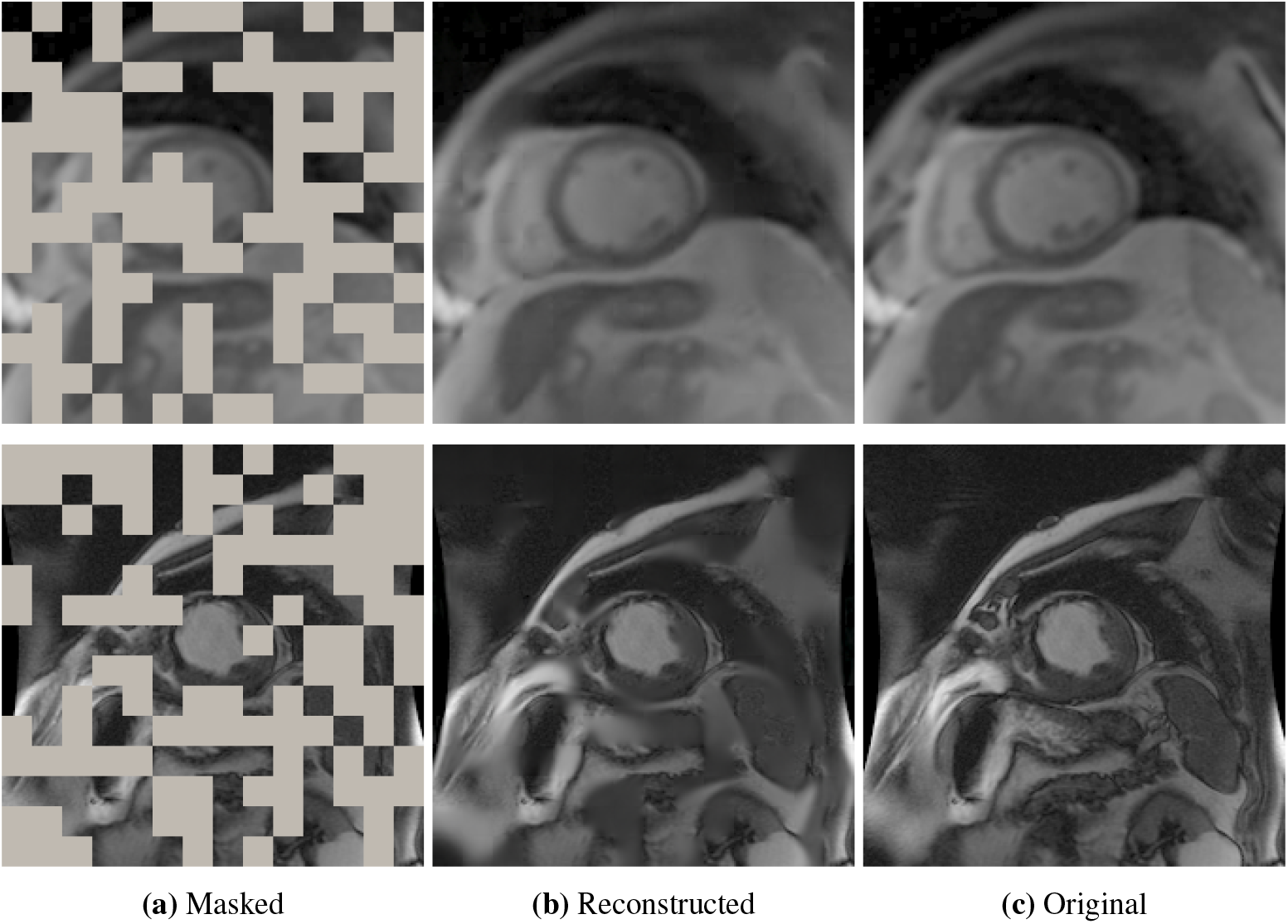
Example MAE reconstruction results demonstrating preservation of ventricular geometry and myocardial contrast across different cardiac phases. For each triplet, we show the the masked image (left), our MAE reconstruction (middle), and the ground-truth (right).

While reconstruction fidelity is not the downstream objective, the ability to recover coherent cardiac structure indicates that the learned representations encode anatomically meaningful information. Such structure-aware embeddings may be particularly beneficial for oxygen saturation prediction in T2-prepared MR images, where signal characteristics within the ventricular blood pool are physiologically informative. These observations support the role of MAE pretraining in learning transferable cardiac representations beyond simple pixel-level reconstruction.

#### 4.4.2 Pooling Strategy Ablation

Since T2-based oximetry acquires multiple images per patient with varying inter-echo spacings, a pooling strategy is required to aggregate per-image features into a single patient-level representation. We investigate pooling strategies for aggregating features across six T2-weighted images per patient, with results shown in Figure 5. Average pooling (mean) generally outperforms max pooling across most configurations, providing balanced representations that incorporate information from all images. From a clinical perspective, averaging across multiple T2-prepared images better reflects the composite physiological state of the blood pool, whereas max pooling can overemphasize isolated frames affected by noise or acquisition variability. This finding supports the use of aggregation strategies that preserve information from all available measurements in multi-echo MR protocols. Additionally, all pre-trained encoders substantially outperform non-pre-trained counterparts regardless of pooling strategy.

**Figure 5:**
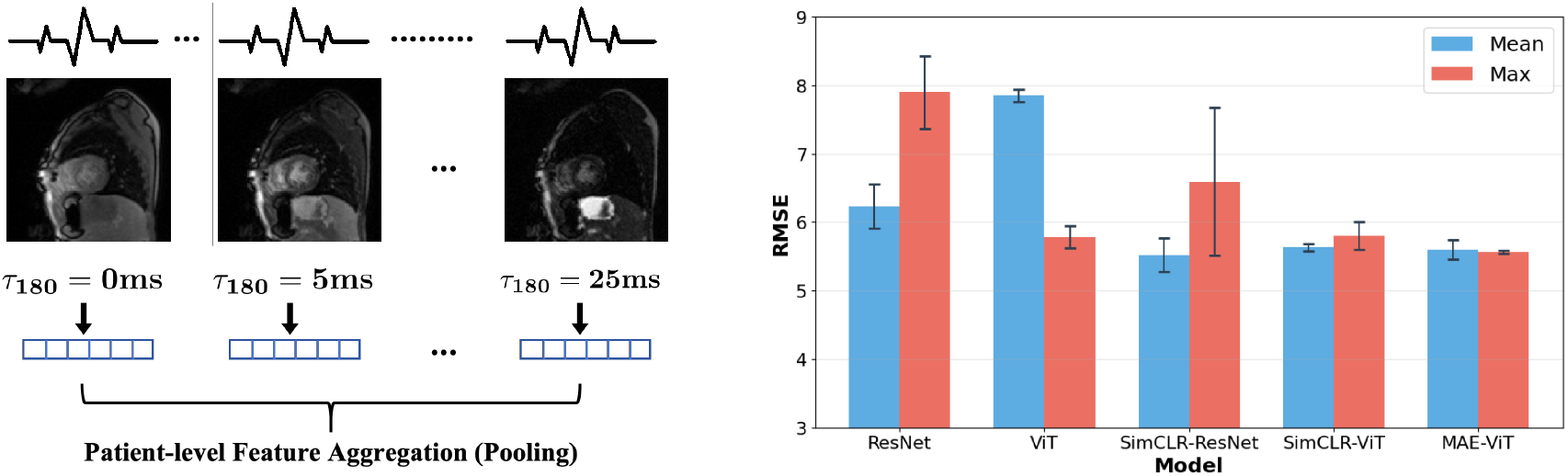
Multi-image feature aggregation for T2 oximetry. (Left) T2-prepared cardiac MR images acquired with varying inter-echo spacings are individually encoded into feature embeddings, which are then aggregated via pooling to form a patient-level representation. (Right) Comparison of pooling strategies (mean vs. max) across different encoder configurations.

#### 4.4.3 Uncertainty Quantification Analysis

Figure 6 shows the relationship between O2 saturation prediction accuracy and estimated uncertainty in two representative examples. High-accuracy cases show narrow Gaussian distributions (high confidence), while larger errors correspond to wider distributions (appropriate low confidence). Attention visualizations reveal that high-confidence predictions focus on specific cardiac regions with sharp attention patterns, while low-confidence cases exhibit diffuse attention with wider uncertainty distributions. This provides clinically meaningful confidence indicators for decision-making. To quantitatively validate this observation, we analyzed the relationship between predicted uncertainty and absolute prediction error across all patients. A strong positive correlation was observed (**r** = **0.83**), indicating that higher uncertainty estimates consistently correspond to larger prediction errors. This suggests that the model appropriately assigns lower confidence to more difficult cases, supporting the potential use of uncertainty as a clinically actionable risk indicator.

**Figure 6:**
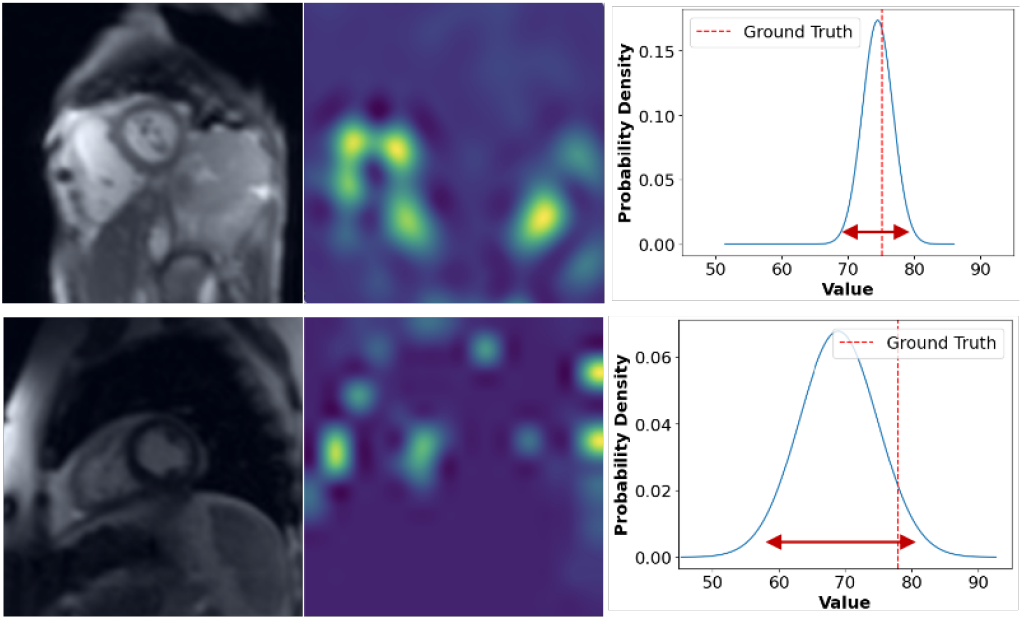
Two representative MR oximetry cases with their attention visualizations and predicted distribution of results w.r.t labelled ground truth.

## 5 Conclusion and Future Work

In this work, we show that self-supervised learning enables accurate prediction of intracardiac blood O2 saturation from T2-sensitive CMR. Pretraining on large-scale unlabeled cardiac MRI yields models that consistently outperform supervised training from scratch. Integrated uncertainty estimates provide calibrated confidence, improving clinical interpretability. These results highlight the potential of leveraging unlabeled cardiac imaging data to address the annotation scarcity challenge in specialized physiological parameter prediction tasks. Future work will focus on validating the proposed framework on larger multi-center cohorts to assess generalizability across diverse patient populations, scanners, and imaging protocols. As larger annotated datasets become available, we plan to explore more advanced multi-image fusion strategies, including attention-based aggregation and transformer-based modeling of T2-prepared image sequences, to better capture complementary physiological information across acquisition settings. Finally, integrating imaging representations with longitudinal clinical data and patient profiles may enable more comprehensive characterization of cardiovascular physiology and support personalized risk assessment and clinical decision-making.

## Data Availability

All data produced in the present study are available upon reasonable request to the authors

## 6 Acknowledgments

This work was supported, in part, by the National Insitute of Biomedical Imaging and Bioengineering under grant number R21EB030294 and the National Science Foundation under award number IIS-2145625. The content is solely the responsibility of the authors and does not necessarily represent the official views of the National Institutes of Health. The authors would like to acknowledge the MR oximetry data contributions of Lajja Desai, MD (Lurie Children’s Hospital), Michael Markl, PhD (Northwestern University), Kevin K. Whitehead, MD, PhD, Matthew A. Harris, MD, and Mark A. Fogel, MD (Children’s Hospital of Philadelphia), Adam Christopher, MD and Laura Olivieri, MD (Children’s National Medical Center), Aimee Armstrong, MD and Kan N Hor, MD (Nationwide Children’s Hospital), Subha V. Raman, MD, Rizwan Ahmad, PhD and Orlando P. Simonetti, PhD (The Ohio State University).

